# Ivermectin for Treatment of Mild-to-Moderate COVID-19 in the Outpatient Setting: A Decentralized, Placebo-controlled, Randomized, Platform Clinical Trial

**DOI:** 10.1101/2022.06.10.22276252

**Authors:** Accelerating COVID-19 Therapeutic Interventions and Vaccines (ACTIV)-6 Study Group, Susanna Naggie

## Abstract

**Background:** The effectiveness of ivermectin to shorten symptom duration or prevent hospitalization among outpatients in the United States with mild-to-moderate symptomatic coronavirus disease 2019 (COVID-19) is unknown.

**Objective:** We evaluated the efficacy of ivermectin 400 µg/kg daily for 3 days compared with placebo for the treatment of early mild-to-moderate COVID-19.

**Methods:** ACTIV-6 is an ongoing, decentralized, double-blind, randomized, placebo-controlled platform trial to evaluate repurposed therapies in outpatients with mild-to-moderate COVID-19. Non-hospitalized adults age ≥30 years with confirmed COVID-19, experiencing ≥2 symptoms of acute infection for ≤7 days were randomized to receive ivermectin 400 µg/kg daily for 3 days or placebo. The main outcome measure was time to sustained recovery, defined as achieving at least 3 consecutive days without symptoms. Secondary outcomes included a composite of hospitalization or death by day 28.

**Results:** Of the 3457 participants who consented to be evaluated for inclusion in the ivermectin arm, 1591 were eligible for this study arm, randomized to receive ivermectin 400 µg/kg (n=817) or placebo (n=774), and received study drug. Of those enrolled, 47% reported receiving at least 2 doses of SARS-CoV-2 vaccination. The posterior probability for any improvement in time to recovery was 0.91 (hazard ratio 1.07, 95% credible interval 0.96–1.17). The posterior probability of this benefit exceeding 24 hours was less than 0.01, as measured by the difference in mean time unwell. Hospitalizations or deaths were uncommon (ivermectin [n=10]; placebo [n=9]). Ivermectin at 400 µg/kg was safe and without serious adverse events as compared with placebo (ivermectin [n=10]; placebo [n=9]).

**Conclusions:** Ivermectin dosed at 400 µg/kg daily for 3 days resulted in less than one day of shortening of symptoms and did not lower incidence of hospitalization or death among outpatients with COVID-19 in the United States during the delta and omicron variant time periods.

**Trial registration:** ClinicalTrials.gov Identifier: NCT04885530.

## INTRODUCTION

There have been advances in the treatment and prevention of coronavirus disease 2019 (COVID-19), caused by severe acute respiratory syndrome coronavirus 2 (SARS-CoV-2). However, there remains a need for additional therapies, particularly in the outpatient setting. Lack of access to or acceptance of vaccines and booster doses has resulted in many people globally remaining at risk of severe COVID-19. Emergence of novel SARS-CoV-2 variants and subvariants has limited the durability of effective monoclonal antibodies, increasing the complexity of clinical care. Use of novel oral antivirals has been authorized to a limited degree in high-income countries,^1,2^ yet implementation challenges exist, potentially due to the limited scope of approval and challenges with drug-drug interactions. There is also a lack of efficacy data for these drugs in people infected with SARS-CoV-2 who have been vaccinated. Currently, in the United States, for those not considered to be at high risk, no targeted therapy is recommended.

Numerous repurposed drugs have been investigated for potential preventative and therapeutic effects in COVID-19.^3-6^ To date, the study of repurposed drugs has been largely in the inpatient setting for the treatment of severe COVID-19.^7-9^ In the outpatient setting, repurposed drug studies have been challenged by small sample sizes, designs with significant limitations, and variable results, limiting the impact on clinical practice.

Ivermectin, an anti-parasitic drug used worldwide for the treatment of onchocerciasis and strongyloidiasis, emerged in 2020 as a potential repurposed drug for COVID-19 due to an *in vitro* study suggesting possible anti-viral activity occurring at concentrations >50-fold higher than achievable in human plasma.^10^ Numerous studies of ivermectin have been completed or are ongoing across multiple populations and the spectrum of COVID-19 disease severity.^11^ While early studies, particularly in the inpatient setting, suggested potential treatment effect, variability in dosing and overall study quality, followed by multiple manuscript retractions, has resulted in controversy.^12-15^ The largest randomized trial to date, the TOGETHER trial, enrolled 1358 patients in Brazil with symptomatic mild-to-moderate COVID-19 and at least 1 risk factor for disease progression. Patients received ivermectin 400 µg/kg for 3 days or placebo in an outpatient setting. No statistically significant clinical benefit of ivermectin was observed for preventing disease progression resulting in hospitalization or prolonged emergency department observation.^16^

ACTIV-6 is an ongoing, fully remote (decentralized), double-blind, randomized, placebo-controlled, platform trial investigating repurposed drugs for the treatment of mild-to-moderate COVID-19 in the outpatient setting. ACTIV-6 is enrolling a broad patient population age ≥30 years in the United States, regardless of comorbid risk factors and vaccine status. Here we report the efficacy of ivermectin 400 µg/kg daily for 3 days compared with placebo for the treatment of early mild-to-moderate COVID-19.

## METHODS

### Trial Design and Oversight

Accelerating COVID-19 Therapeutic Interventions and Vaccines (ACTIV)-6 (NCT04885530) is a double-blind, randomized, placebo-controlled platform protocol designed to be flexible, allowing for use in a wide range of settings within healthcare systems and the community for integration into routine COVID-19 testing programs and subsequent treatment plans. The platform protocol enrolls outpatients with mild-to-moderate COVID-19 with a confirmed positive polymerase chain reaction (PCR) or antigen test for SARS-CoV-2, including home-based testing. Each repurposed medication (study drug arm/group) is further described including drug-specific exclusion criteria in each drug-specific appendix.

A governing institutional review board for each site approved the protocol. Informed consent was obtained from each enrolled participant either via e-consent or (in person consent) written process. An independent data monitoring committee oversaw the monitoring of participant safety, efficacy, and trial conduct.

### Participants

The platform trial opened recruitment on June 11, 2021, and is ongoing. Participants were enrolled into the ivermectin arm or identical matched-placebo or contributing-placebo from June 23, 2021 through February 4, 2022 at 93 sites in the United States. Participants were either identified by sites or self-identified by contacting central study telephone hotline(s).

Sites verified eligibility criteria including age ≥30 years, confirmed SARS-CoV-2 infection within 10 days, and experiencing ≥2 symptoms of acute COVID-19 for ≤7 days from enrollment. Symptoms included fatigue, dyspnea, fever, cough, nausea, vomiting, diarrhea, body aches, chills, headache, sore throat, nasal symptoms, and loss of sense of taste or smell. Exclusion criteria included hospitalization, study drug use within 14 days, or known allergy or contraindication to study drug. Ivermectin-specific exclusion criteria were end-stage kidney disease on renal replacement therapy, liver failure, decompensated cirrhosis, pregnancy, or breastfeeding. Vaccination was allowable, as were standard of care therapies for COVID-19 provided under United States Food and Drug Administration (FDA)-approval or emergency use authorization (EUA).

### Randomization and Interventions

Participants were randomized in a two-step process. First, each participant was randomized with equal probability among the study drugs actively enrolling for which the participant was eligible. Participants could choose to opt out of specific study drugs if they or the site investigator did not feel there was equipoise. Each participant was required to select at least one study drug and may have selected all active study drugs available on the platform at the time of enrollment.

Subsequent to randomization among study drugs, participants were randomized to either active agent or to placebo in a ratio of *m*:1 where *m* is the number of study drugs for which the participant was eligible. The more study arms a participant was eligible for, the greater the chance of receiving an active study drug. All participants randomized to the ivermectin study drug are included in the ivermectin arm or the *matched* placebo arm. Participants eligible for the ivermectin study drug but randomized to placebo for a different study drug are included and *contribute* to the placebo arm.

A central pharmacy supplied ivermectin or placebo to participants via direct home delivery. Ivermectin was supplied as a bottle of fifteen 7-mg tablets. Participants were instructed to take a pre-specified number of tablets for 3 consecutive days based on their weight for a daily dose of approximately 400 µg/kg. Packaging for matched placebo was identical to ivermectin. Packaging for other contributing placebo was identical to the associated study drug.

### Outcome Measures

The primary measure of effectiveness was based on time to sustained recovery, defined as achieving at least three consecutive days without symptoms; this was selected *a priori* from among the two co-primary endpoints that remain available to other study drugs in the platform. The key secondary outcome is a composite of hospitalization or death by day 28. Other secondary outcomes include the COVID Clinical Progression Scale on days 7, 14, and 28; mortality through day 28; and hospitalization, urgent care visit, or emergency department visit through day 28.

### Trial Procedures

ACTIV-6 is designed as a fully remote, or decentralized, trial. All study visits are designed to be remote. Screening and eligibility confirmation were participant-reported and site confirmed. A positive SARS-CoV-2 PCR or antigen test result was verified prior to randomization. At screening, demographic information, eligibility criteria, medical history, concomitant medications, symptom reporting, and quality of life questionnaires were participant-reported. However, screening and enrollment could occur in-person at sites and unplanned study visits could occur in-person or remotely, as deemed appropriate by the site investigator.

For distribution of study drug, a central investigational pharmacy was used. Participants were required to provide a valid residential address at which they could receive a package. Shipping and delivery were tracked. Participants must receive study drug to be included in the analysis; receipt of study drug is defined as Day 1 for this study.

Participants were asked to complete daily assessments and report safety events via the study portal through day 14, then at other intervals through day 28, and at the final study visit at day 90. Assessments included symptoms and severity, health care visits, and medications. If participants were still reporting symptoms at day 14, they continued to be assessed until they experienced three consecutive days without symptoms or until day 28. At days 28 and 90, all participants completed assessments.

The daily and follow-up assessments were monitored, and sites were actively notified of events requiring review, including serious adverse events. In addition, participants were invited during these assessments to request contact from the study team or to report any unusual circumstances that might be relevant. Failure to complete daily assessments was also a trigger for review of a possible serious adverse event. A missed assessment on the day after receiving the first dose of study medication (day 2) or any day of missed assessments up to day 14 prompted an investigator notification to contact the participant. All participants were instructed to self-report concerns either via an online event reporting system, by calling the site, or by calling a 24-hour hotline.

Hospitalizations, a record of seeking other healthcare, or serious adverse events were extracted by site personnel from the participant’s medical record. Medical occurrences occurring before the receipt of study drug/placebo but after obtaining informed consent were not considered an adverse event.

### Independent Data Monitoring Committee Oversight

Due to extremely rapid enrollment related to the omicron variant surge, 2000 participants were enrolled in the platform trial from December 15, 2021 to February 1, 2022. This resulted in the full accrual of the ivermectin arm before the first planned interim analysis by the independent data monitoring committee review.

### Statistical Analysis Plan

This ongoing platform trial is designed to be analyzed accepting the possibility of adding and dropping arms as the trial progresses. The general analytical approach is Bayesian regression modelling. Proportional hazard regression was used for time-to-event analysis, and cumulative probability ordinal regression models were used for ordinal outcomes. In addition, the mean time spent unwell was estimated using a longitudinal ordinal regression model as a quantification of benefit. Interim analyses are planned at intervals of approximately 300 participants contributing to a study drug arm. There is also the potential for extending accrual for a study drug if there is the potential to demonstrate benefit for hospitalization/death. Decision thresholds were set to balance overall power with control of the Type I error rate in the context of the study drug-specific objectives. A sample size of approximately 1200 participants per study drug was determined to be sufficient to conclude whether there is meaningful evidence of benefit based on a reduction in time to sustained recovery.

As a platform trial, the primary analysis is implemented separately for each study drug, where the placebo arm consists of concurrently randomized participants that meet the eligibility criteria for that study drug; this includes both matched and contributing placebo. From other remote trials,^3,6^ it was recognized that medication delivery (placebo or study drug) may not always occur (e.g., failure of delivery, participant withdrawal, or interval hospitalization). For this trial, a modified intention-to-treat approach was specified for the primary analyses to include all participants who received study drug. All available data were used to compare each study drug versus placebo control, regardless of post-randomization adherence to study protocols.

A pre-specified analysis tested for differential treatment effects as a function of pre-existing participant characteristics. Analysis of heterogeneity of treatment effect included age, number of days of symptoms, body mass index, symptom severity, calendar time (surrogate for SARS-CoV-2 variant), and vaccination status; continuous variables were modelled as such without creating subgroups.

## RESULTS

### Study Population

Of the 3457 participants who consented to be evaluated for inclusion in the ivermectin arm, 1591 were eligible for this study arm, randomized to ivermectin 400 µg/kg (n=817) or placebo (n=774), and received study drug (**eFigure 1**). Of participants receiving placebo, 545 (70%) received matching placebo and 229 (30%) received placebo as part of a concurrent study arm and contributed to the pooled placebo group.

The mean age of the participants was 48 years (SD 12), and 43% were aged 50 years or older (**Table 1**). The population was 59% female, 81% identified as White, 7% identified as Black/African American, and 10% reported being of Latino/Hispanic ethnicity. Although not required for enrollment, high-risk comorbidities were prevalent, including body mass index >30 kg/m^2^ (41%), diabetes (11.5%), hypertension (26%), asthma (15%), and chronic obstructive pulmonary disease (4%). Vaccination was common, with 47% reporting having received at least 2 doses of vaccine. The median time from symptom onset to receipt of study drug was 6 days (IQR 4 to 8). Baseline symptom prevalence and severity are described in **eTable 1**. Use of therapies available under FDA-approval or authorization was uncommon (remdesivir 0.3%, monoclonal antibody 3%, ritonavir boosted nirmatrelvir 0.1%).

**Table 1.**
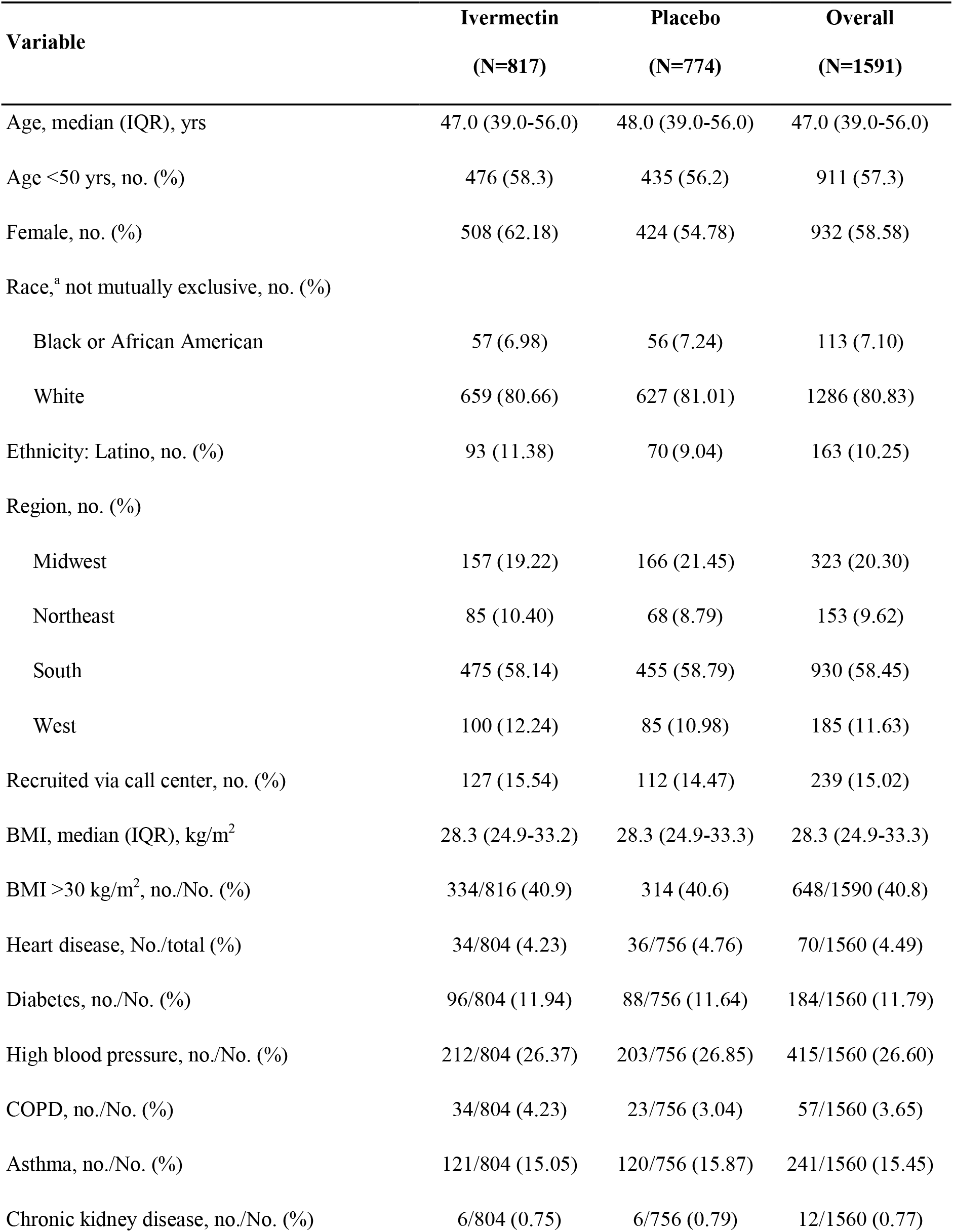
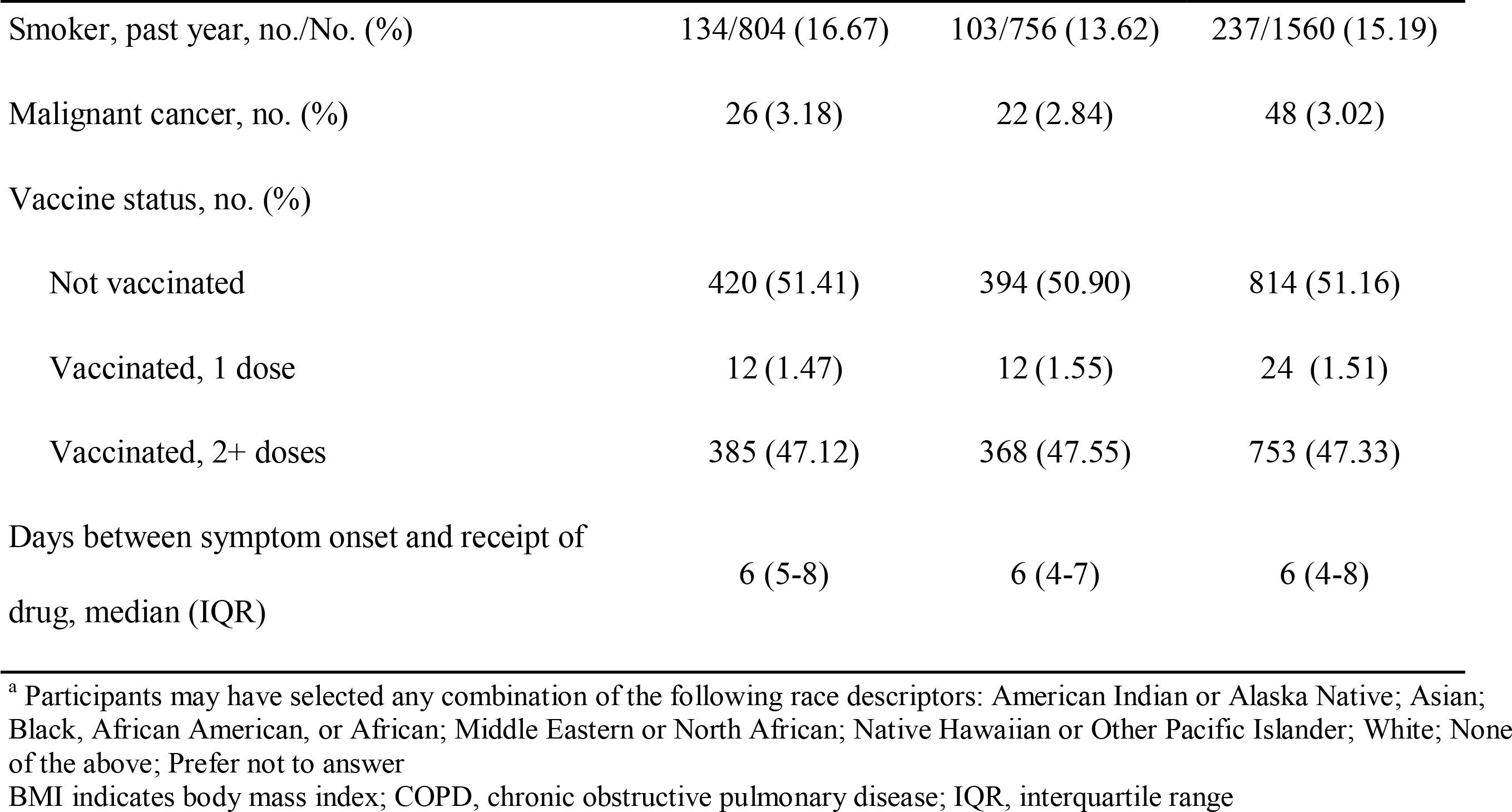
Baseline characteristics

| **Variable** | **Ivermectin**<br>**(N=817)** | **Placebo**<br>**(N=774)** | **Overall**<br>**(N=1591)** |
| --- | --- | --- | --- |
| Age, median (IQR), yrs | 47.0 (39.0-56.0) | 48.0 (39.0-56.0) | 47.0 (39.0-56.0) |
| Age <50 yrs, no. (%) | 476 (58.3) | 435 (56.2) | 911 (57.3) |
| Female, no. (%) | 508 (62.18) | 424 (54.78) | 932 (58.58) |
| Race,<sup>a</sup> not mutually exclusive, no. (%) |  |  |  |
| Black or African American | 57 (6.98) | 56 (7.24) | 113 (7.10) |
| White | 659 (80.66) | 627 (81.01) | 1286 (80.83) |
| Ethnicity: Latino, no. (%) | 93 (11.38) | 70 (9.04) | 163 (10.25) |
| Region, no. (%) |  |  |  |
| Midwest | 157 (19.22) | 166 (21.45) | 323 (20.30) |
| Northeast | 85 (10.40) | 68 (8.79) | 153 (9.62) |
| South | 475 (58.14) | 455 (58.79) | 930 (58.45) |
| West | 100 (12.24) | 85 (10.98) | 185 (11.63) |
| Recruited via call center, no. (%) | 127 (15.54) | 112 (14.47) | 239 (15.02) |
| BMI, median (IQR), kg/m<sup>2</sup> | 28.3 (24.9-33.2) | 28.3 (24.9-33.3) | 28.3 (24.9-33.3) |
| BMI >30 kg/m<sup>2</sup>, no./No. (%) | 334/816 (40.9) | 314 (40.6) | 648/1590 (40.8) |
| Heart disease, No./total (%) | 34/804 (4.23) | 36/756 (4.76) | 70/1560 (4.49) |
| Diabetes, no./No. (%) | 96/804 (11.94) | 88/756 (11.64) | 184/1560 (11.79) |
| High blood pressure, no./No. (%) | 212/804 (26.37) | 203/756 (26.85) | 415/1560 (26.60) |
| COPD, no./No. (%) | 34/804 (4.23) | 23/756 (3.04) | 57/1560 (3.65) |
| Asthma, no./No. (%) | 121/804 (15.05) | 120/756 (15.87) | 241/1560 (15.45) |
| Chronic kidney disease, no./No. (%) | 6/804 (0.75) | 6/756 (0.79) | 12/1560 (0.77) |
| Smoker, past year, no./No. (%) | 134/804 (16.67) | 103/756 (13.62) | 237/1560 (15.19) |
| Malignant cancer, no. (%) | 26 (3.18) | 22 (2.84) | 48 (3.02) |
| Vaccine status, no. (%) |  |  |  |
| Not vaccinated | 420 (51.41) | 394 (50.90) | 814 (51.16) |
| Vaccinated, 1 dose | 12 (1.47) | 12 (1.55) | 24 (1.51) |
| Vaccinated, 2+ doses | 385 (47.12) | 368 (47.55) | 753 (47.33) |
| Days between symptom onset and receipt of<br>drug, median (IQR) | 6 (5-8) | 6 (4-7) | 6 (4-8) |

<sup>a</sup> Participants may have selected any combination of the following race descriptors: American Indian or Alaska Native; Asian; Black, African American, or African; Middle Eastern or North African; Native Hawaiian or Other Pacific Islander; White; None of the above; Prefer not to answer
BMI indicates body mass index; COPD, chronic obstructive pulmonary disease; IQR, interquartile range

### Primary and Secondary Outcomes

In the modified intention-to-treat population, we observed that the posterior probability for benefit on the primary outcome of time to recovery between the ivermectin and placebo arms was 0.91 (hazard ratio [HR] 1.07, 95% credible interval [CrI] 0.96 to 1.17) where a HR >1 is for faster symptom resolution (**Table 2, Figure 1**). The difference in the amount of time spent feeling unwell with COVID-19 was estimated to be 0.49 days (95% CrI, 0.15 to 0.82 days) in favor of ivermectin. The posterior probability that this benefit exceeds one day is less than 0.01 (**Figure 2A**). There were no differences in secondary outcomes (**Figure 2B-C**). Hospitalization or death were uncommon, occurring in 1.22% (10/817) with ivermectin and 1.16% (9/774) with placebo; there was one death in the ivermectin arm (**Table 2, eFigure 2A**). Similarly, the composite secondary outcome of urgent or emergency care visits, hospitalizations, or death were similar with ivermectin (3.9% [32/817]) compared with placebo (3.6% [28/774]) (**Table 2, eFigure 2B**). The posterior probability for treatment benefit did not meet prespecified thresholds for clinical events or on the COVID Clinical Progression Scale (**Online Supplement**) at days 7, 14, and 28 (**Table 2**).

**Figure 1.**
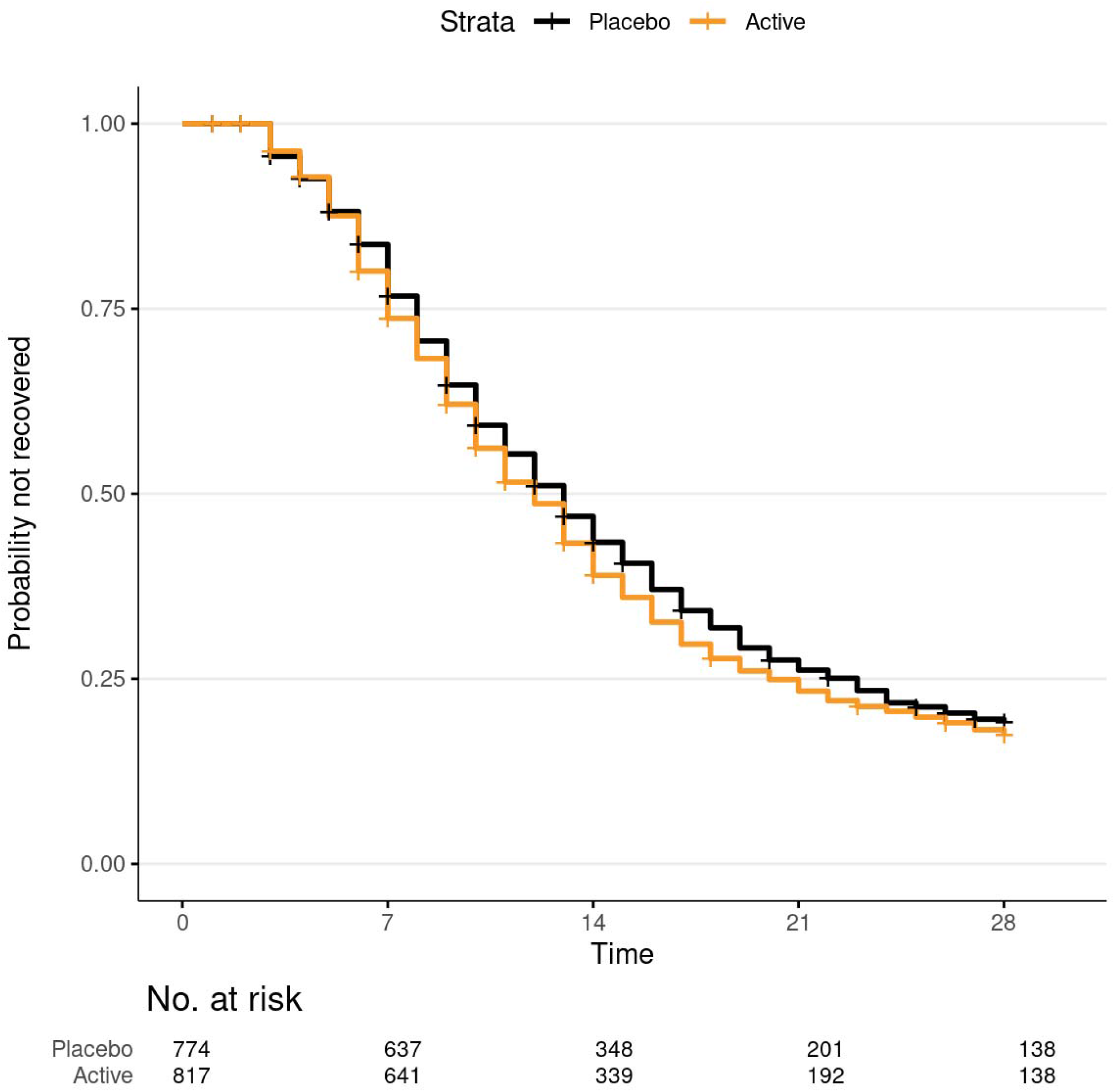
Time to recovery from COVID-19 with ivermectin versus placebo. Kaplan-Meier curve and 95% confidence intervals (point-wise) for time-to-recovery endpoint. The mean days unwell was 10.96 with ivermectin versus 11.45 with placebo. Among participants that did not die during follow-up, recovery was defined as three consecutive days without COVID-19 symptoms, as affirmatively reported by the study participant. Time to sustained recovery was the number of days between the receipt of study drug and achieving three consecutive days without symptoms. Participants that died, by definition, did not recover regardless of reported symptom freedom. Time to recovery was administratively censored at 28 days.

**Figure 2A.**
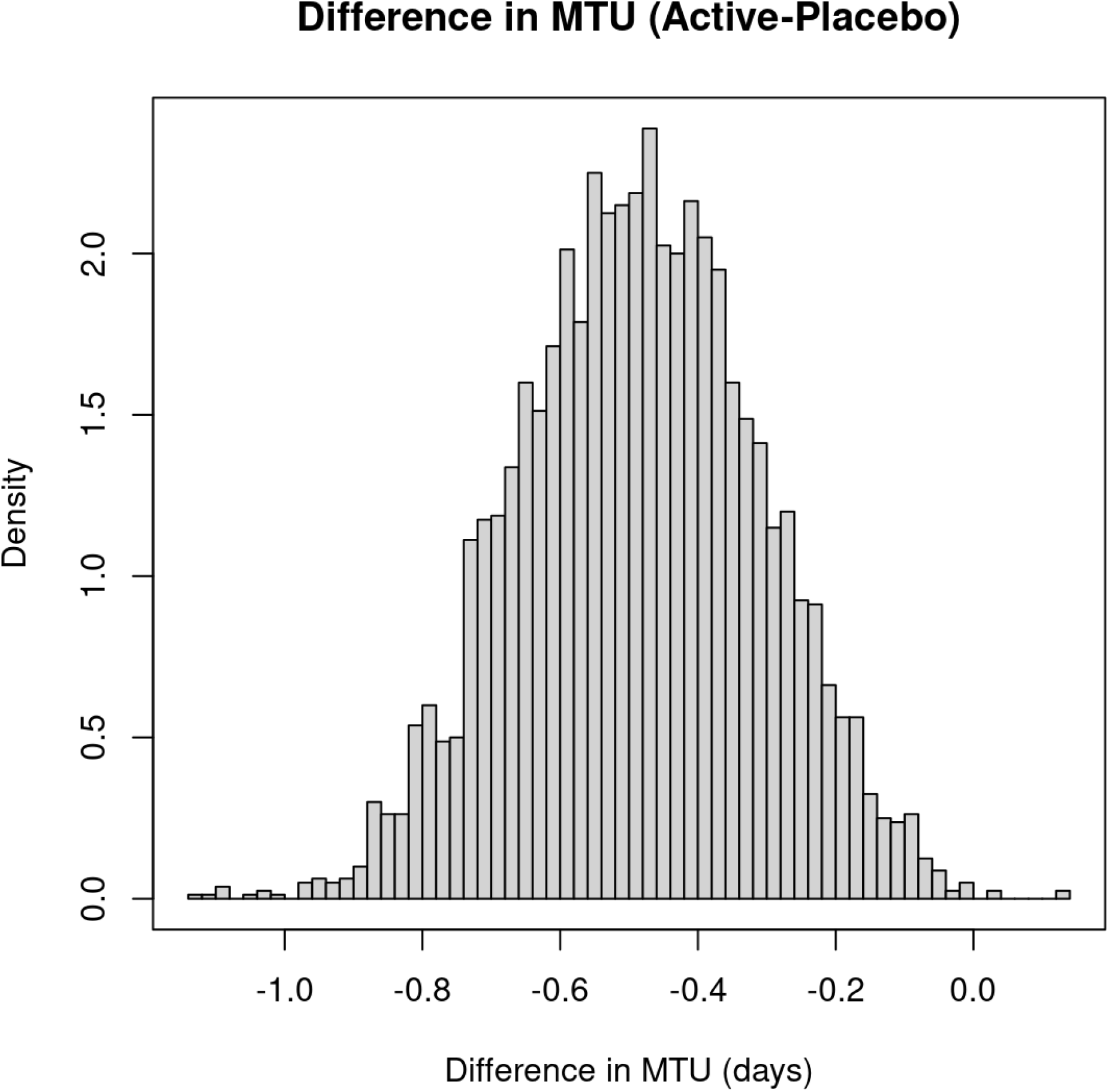
Posterior distribution of the difference in mean time unwell (MTU). Mean time unwell is a model-based estimate of the number of days with symptoms or hospitalized or deceased during the first 14 days of follow-up. Negative differences in MTU (Active-Placebo) indicate that participants in the placebo arm were unwell longer than participants in the ivermectin arm. The estimate of MTU is calculated from a Bayesian, longitudinal, ordinal regression model with covariates age (as restricted cubic spline) and calendar time. The prior distribution was not informative. The difference in MTU was -0.49 (95% CrI: -0.82, -0.15). The posterior probability that the difference was larger than 1 day was less than 0.01.

**Figure 2B.**
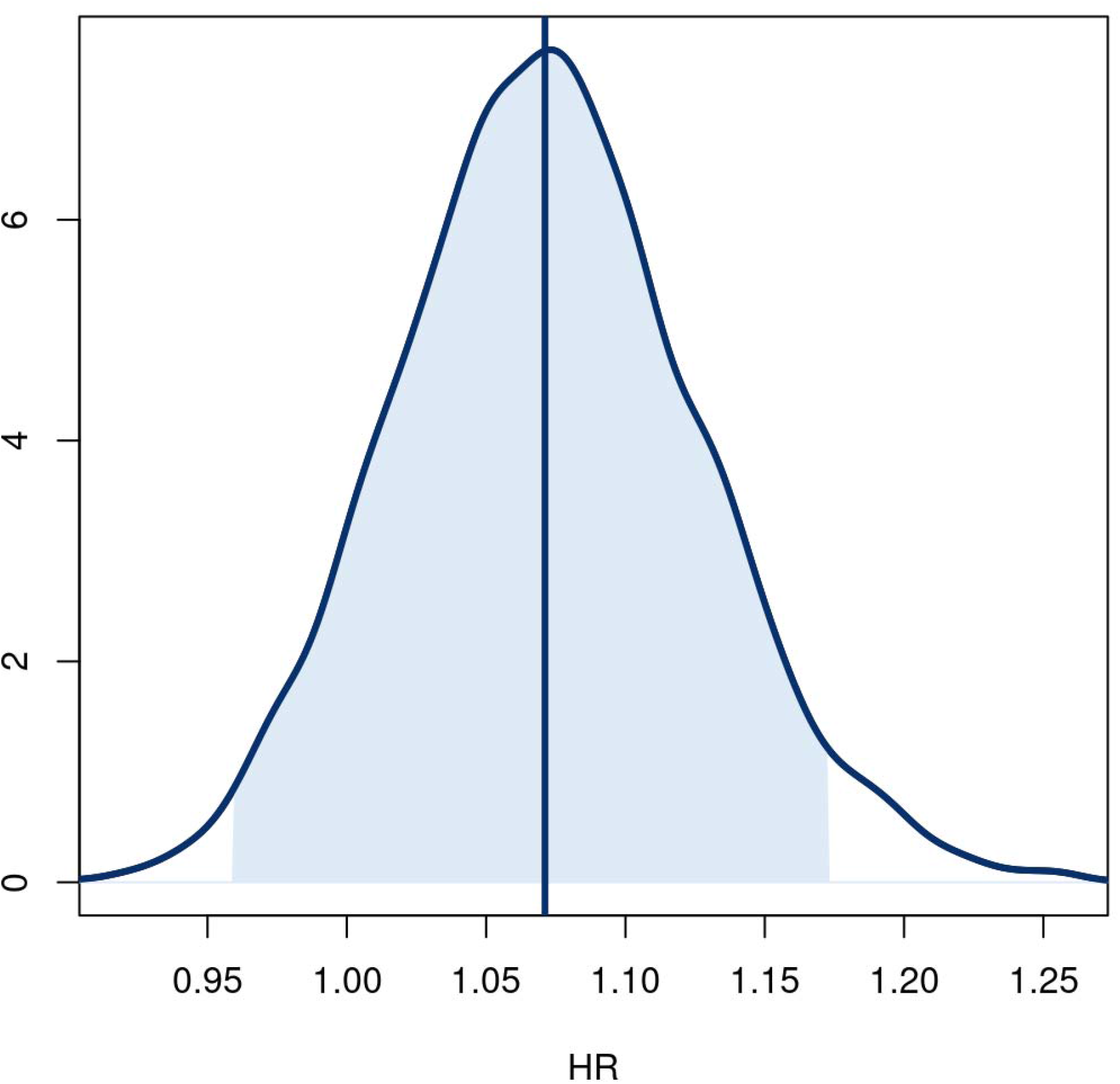
Posterior distributions of treatment effect hazard ratio for time to sustained recovery. Posterior distribution for the treatment effect hazard ratio for the time to sustained recovery. The posterior was based on a covariate adjusted Cox proportional hazards regression with skeptical prior. The baseline hazard was a degree 5 M-spline function. Covariates in the model included age (as restricted cubic spline), sex, duration of symptoms, vaccine status, geographic region, origination from call center, calendar time (as restricted cubic spline), and symptom burden on the day of drug receipt. Hazard ratios greater than 1.0 favor ivermectin.

**Figure 2C.**
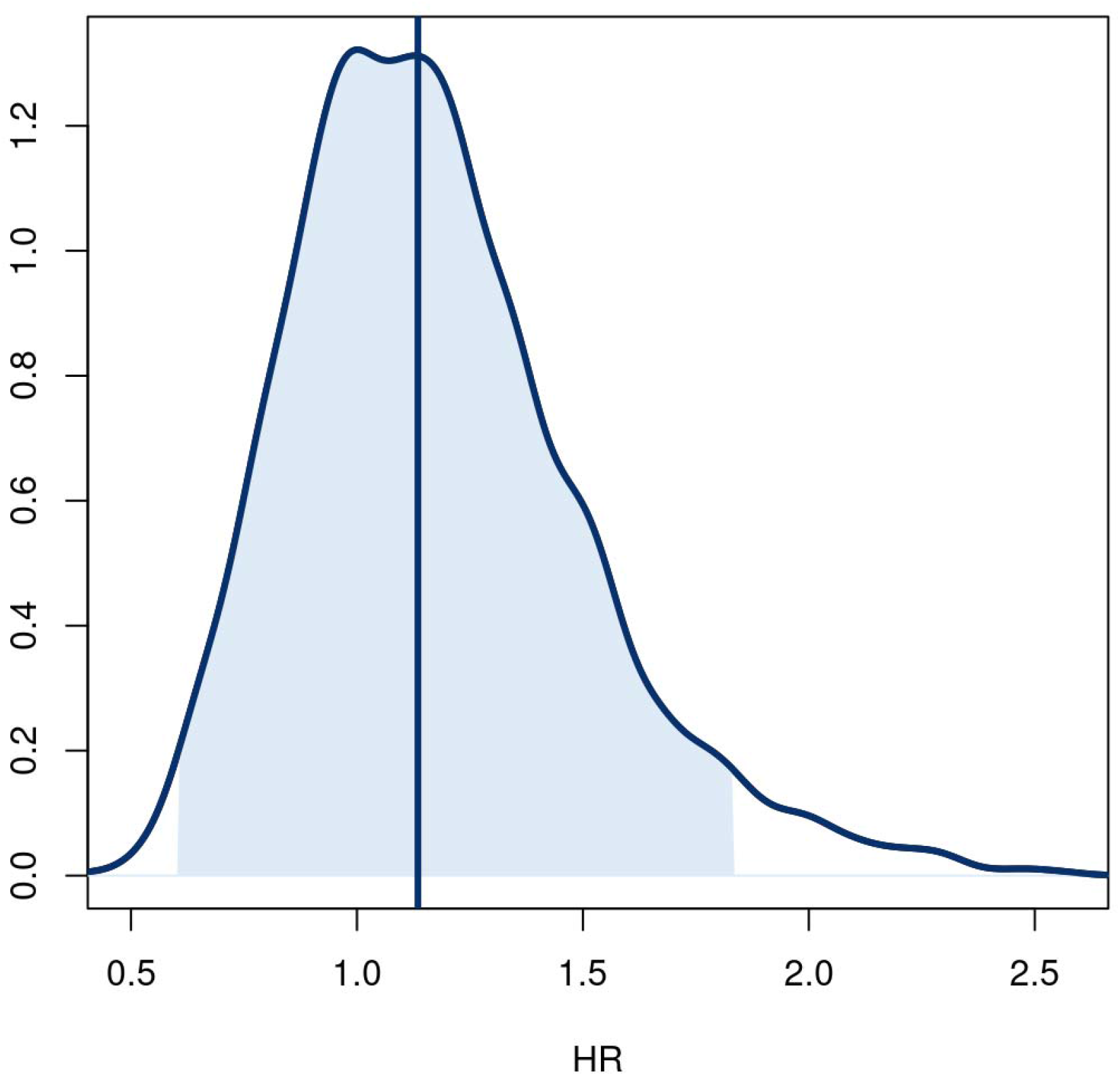
Posterior distributions of treatment effect hazard ratio for time to the composite endpoint of hospitalization, urgent care, emergency room visit, or death through day 28. Posterior distribution for the treatment effect hazard ratio for the time to the composite endpoint of hospitalization, urgent care, emergency room visit, or death through day 28. The posterior was based on a covariate adjusted Cox proportional hazards regression with uninformative prior. The baseline hazard was a degree 5 M-spline function. Covariates in the model included age (as restricted cubic spline), sex, duration of symptoms, vaccine status, geographic region, origination from call center, calendar time (as restricted cubic spline), and symptom burden on the day of drug receipt. Hazard ratios less than 1.0 favor ivermectin.

**Table 2.** Primary and secondary outcomes

|  | **Ivermectin** | **Placebo** | **Estimate** | **Posterior** |
| --- | --- | --- | --- | --- |
|  | **(N=817)** | **(N=774)** | **(95% Interval)<sup>a</sup>** | **P(efficacy)** |
| Time to recovery |  |  |  |  |
| Skeptical prior (primary) |  |  | HR: 1.07 (0.96, 1.17) | 0.91 |
| Non-informative prior |  |  | HR: 1.09 (0.97, 1.22) | 0.93 |
| No prior |  |  | HR: 1.09 (0.98, 1.22)<sup>b</sup> | - |
| Mortality at day 28, No. (%) | 1 (0.12) | 0 (0.00) | - | - |
| Hospitalization or death through day 28, No. (%) | 10 (1.22) | 9 (1.16) | HR: 1.1 (0.4, 2.6)<sup>b, c</sup> | -<sup>d</sup> |
| Hospitalization, urgent care, emergency room visit, or death through day 28, No. (%) | 32 (3.9) | 28 (3.6) | HR: 1.2 (0.6, 1.8) | 0.32 |
| Clinical progression ordinal outcome scale |  |  |  |  |
| Day 7 |  |  | OR: 0.76 (0.55, 1.00) | 0.97 |
| Day 14 |  |  | OR: 0.73 (0.52, 0.98) | 0.98 |
| Day 28 |  |  | OR: 0.90 (0.60, 1.21) | 0.74 |
| Mean time unwell, days (95% CrI)<sup>e</sup> | 10.96 (10.78, 11.15) | 11.45 (11.28, 11.60) | $\Delta$: -0.49 (-0.82, -0.15) | 0.99 |

<sup>a</sup> Unless otherwise noted, a highest density credible interval. <sup>b</sup> Confidence interval. <sup>c</sup> Low event rate precluded covariate adjustment. <sup>d</sup> Due to the low event rate, a posterior probability was not estimated. <sup>e</sup> The mean time unwell is estimated from receipt of study drug to achieving sustained recovery. For direct comparison to studies that use the first day of recovery, two days should be subtracted from these estimates
Abbreviations: HR = hazard ratio; OR = Odds Ratio
Time to Recovery, HR >1.0 is favorable for faster recovery; Clinical progression ordinal outcome scale OR<1.0 is favorable for less progression

### Heterogeneity of treatment effect analyses

Tests for heterogeneity of treatment effect show no overall influence of the putative subgrouping variables on treatment effects. While those reporting severe symptoms possibly experienced benefit of faster symptom resolution with ivermectin (HR 1.79; 95% CrI, 1.06 to 3.04), the overall effect of symptom severity was not significant (p=0.123) and the small sample size in the severe group (51 ivermectin vs. 39 placebo) suggests this results should be considered exploratory requiring further validation in a future trial (**Figure 3**). There was no evidence of a different treatment effect with ivermectin compared with placebo for timing of symptom onset, body mass index, calendar time, or vaccination status.

**Figure 3.**
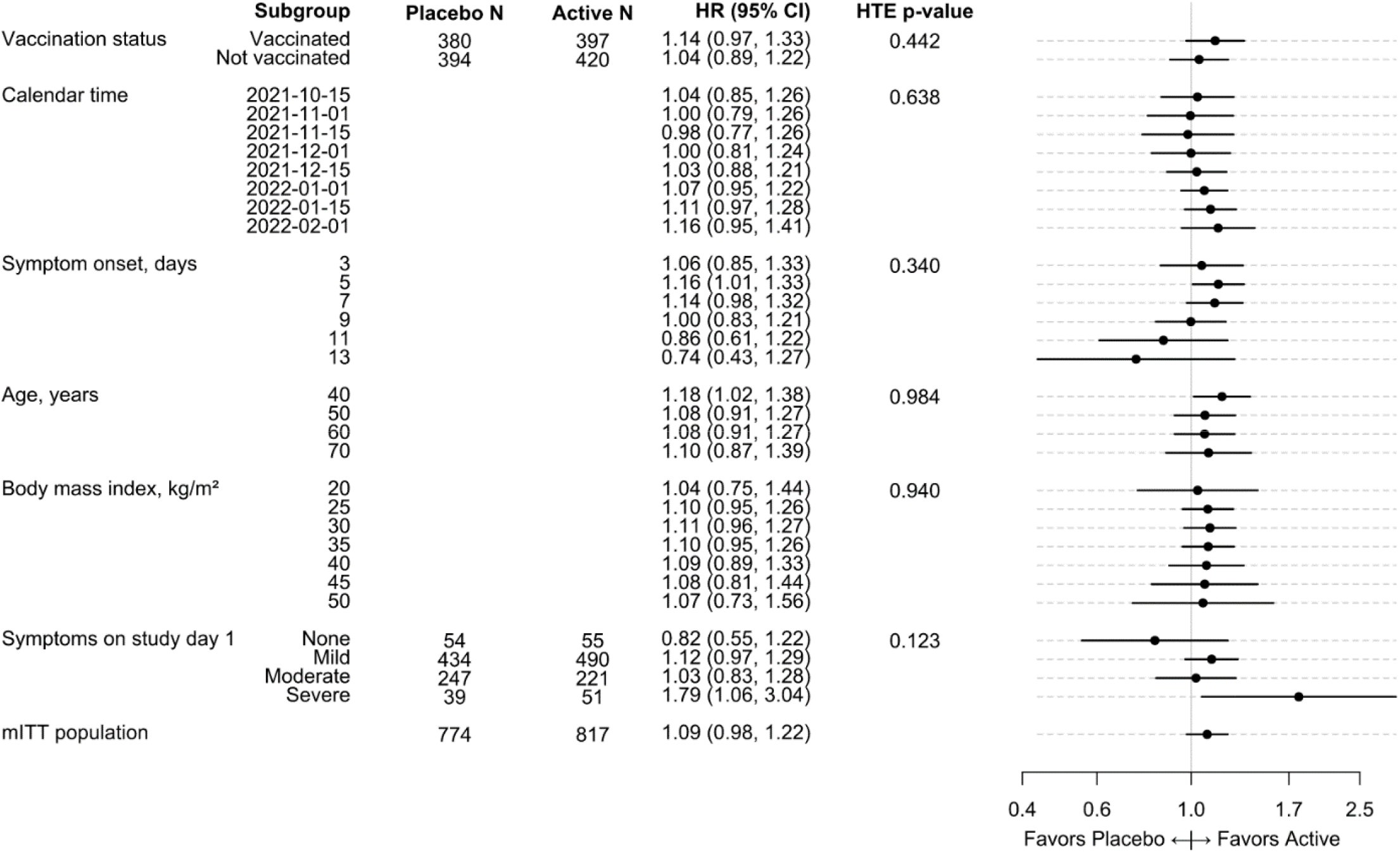
Covariate-adjusted and model-based estimates of the treatment effect for selected characteristics for ivermectin versus placebo. A hazard ratio greater than 1.0 indicates a faster time to recovery. Study day 1 was the day of starting the study medication. The ‘mITT population’ reflects a modified intent-to-treat analysis of participants randomized who received study medicine within 7 days. Covariate-adjusted and model-based estimates of the treatment effect for selected subgroups. For each characteristic, a proportional hazards regression model was constructed using the same covariates as the primary endpoint model plus additional interaction terms between treatment assignment and the characteristic of interest. For example, the interaction of vaccination status and treatment assignment was added to the primary endpoint regression model to calculate a treatment effect for the vaccinated and unvaccinated subgroups. To allow the possibility of non-linear trends along continuous characteristics, like age or calendar time, the additional terms were interactions between treatment assignment and restricted cubic splines. Because the primary endpoint model did not include body mass index (BMI), the restricted cubic spline terms for BMI were also added to the model (sometimes call main effects) in addition to the interaction terms. Because the primary endpoint model only included a single linear term for symptom onset, the nonlinear terms of the restricted cubic spline were also added to the model in addition to the interaction terms. The hazard ratios and 95% confidence intervals were calculated from asymptotic, model-based contrasts. The hazard ratio for the full study population was generated from the primary endpoint model without prior.

### Safety

Adverse events were uncommon and similar in both arms (2.8% with ivermectin; 3.5% with placebo). All but one recorded event occurred in participants who confirmed taking their study drug; one participant who reported not taking study drug experienced acute kidney injury. Ivermectin at 400 µg/kg was safe and without additional serious adverse events compared with placebo (ivermectin [n=10]; placebo [n=9]) (**eTable 2**).

## DISCUSSION

We did not find a clinically relevant effect for treatment of early COVID-19 with ivermectin 400 µg/kg daily for 3 days in this large trial that enrolled over 1500 participants in the United States. The lack of treatment effect was consistent for the primary outcome of time to symptom resolution and the secondary clinical outcomes including hospitalization, death, or acute care visits. While a more sensitive analysis identified a favorable difference for the ivermectin arm in the amount of time spent feeling unwell with COVID-19, the probability that this effect exceeds one day was less than 0.01.

Although there are numerous published studies reporting on the potential efficacy of ivermectin for the treatment of COVID-19, many are in the inpatient setting and the majority are small, variable in population and dosing, and some have been retracted.^12-15^ In the outpatient setting, larger well-designed trials such as ACTIV-6 are emerging and do not support a substantial clinical benefit of ivermectin when used at a dose of 400 µg/kg daily for 3 days. The outcomes studied from these outpatient trials are comprehensive and include symptom resolution, acute healthcare utilization, hospitalization, and death.^16^ Thus, ACTIV-6 adds to the growing evidence that there is not a clinically relevant treatment effect of ivermectin at this dose and duration. While those with severe symptoms at baseline appeared to have beneficial treatment effect with ivermectin as compared with placebo, this subgroup was small, thus these findings should be considered exploratory.

ACTIV-6 has several notable strengths. Many of the prior studies of ivermectin were conducted largely outside of the United States, thus data from a high-income country with associated healthcare system were lacking. ACTIV-6 is a double-blind, randomized, placebo-controlled national study with enrolling sites in 28 states and a call center able to recruit participants from the remainder of the United States. This ivermectin arm of the ACTIV-6 platform trial enrolled rapidly due to the delta and omicron variant surges and included both vaccinated and unvaccinated patients, thus representing a highly relevant study population. The trial also has limitations. Due to the broad study population, including almost 50% reporting vaccination, few participants progressed to severe COVID-19, limiting the power to study the treatment effect on relevant clinical outcomes like hospitalization and death. Due to the remote nature of the trial and constraints related to timing of randomization, the average time from start of symptoms to receipt of study drug was 6 days, which is later in the disease course than recent antiviral trials.^1,2^ However, there was no benefit observed for those who started treatment earlier (≤3 days) versus later (>3 days) in the subgroup analysis.

This large ACTIV-6 trial did not identify a clinically relevant treatment effect with ivermectin 400 µg/kg dosed daily for 3 days; the subgroup analyses and overall totality of data supports further investigation of a higher dose and longer course of ivermectin for the treatment of mild-to-moderate COVID-19 in the outpatient setting.

## Data Availability

Prior to deposition of the data in a public repository which will occur when the platform trial has concluded, persons may request the data by submitting a proposal. If the data can be used for the proposed purpose and it is consistent with the informed consent, then the data will be released under a Data Use Agreement.

## Author Contributions

Author and collaborator contributions, including responsibility for decision to submit the manuscript, drafting of the initial manuscript, study conceptualization, investigation, data curation, formal analysis, study supervision, and review and editing of the manuscript, are provided in the Online Supplement. CL and TS directly accessed and verified the underlying study data. SN and AFH had access to all the study data and had final responsibility for the decision to submit the paper for publication.

